# Predictions of Covid-19 Related Unemployment On Suicide and All-cause Mortality

**DOI:** 10.1101/2020.05.02.20089086

**Authors:** Rajiv Bhatia

## Abstract

**Importance:** The Covid-19 pandemic has driven behavioral and governmental responses with large impacts on economic activity. Estimates of indirect health effects of economic impacts may inform societal action.

**Objective:** To estimate the size of the impact of Covid-19 unemployment on suicides and deaths from all causes.

**Design:** Risk assessment applying of pooled effects hazard ratios from published meta-analyses of observational epidemiological studies, post-Covid-19 unemployment, current labor force composition data, and current age-adjusted mortality rates.

**Results:** This risk assessment estimates approximately 9,700 excess annual deaths from suicide and 66,000 annual deaths from all causes among those recently unemployed due to Covid-19.

**Conclusions and Relevance:** Indirect health impacts of societal responses to Covid-19 are identifiable, multiple and quantifiable. Adverse health impacts, such as those from unemployment, may endure longer than those of the Covid-19 pandemic itself. Decision-makers can include indirect health impacts in policy-making calculi for Covid-19 mitigation and suppression strategies.

**Key Points:** *Question:* What are the expected impacts of post Covid-19 unemployment on excess suicide and premature death.

*Findings:* A risk assessment applying pooled summary risk estimates from meta-analyses of observational studies predicts approximately 9,700 excess annual deaths from suicide and 66,000 annual deaths from all causes among those recently unemployed due to Covid-19.

*Meaning:* Indirect health impacts of societal responses to Covid-19 are identifiable, multiple and quantifiable.

## Introduction

To limit death and mitigate demand on health care systems from the transmission of Covid-19 governments employed rarely used policies to limit mobility and economic activity, including travel restrictions, bans on gathering, and community-wide stay-at-home rules. Uncertainty and health threat required rapid decisions based on non-peer reviewed predictive models and without an explicit trade-offs analysis. (1)

Behavioral responses to Covid-19 re-enforced by governmental social distancing rules have resulted in a number of secondary consequence, business closures, unemployment, bankruptcies, and delayed health care, fear and anxiety, and suicide crisis line calls.

Available risk assessment tools applied to some of these indirect impacts might quantify secondary health effects of these impacts. (2,3) Weighing such indirect health impacts might contribute to balancing responses to control Covid-19 with other health aims and may inform the nature, scope, and adoption of policy countermeasures, such as income supports and expansion of health and mental health services.

Between the onset of the coronavirus pandemic and May 14th, 2020, over 36 million Americans had filed unemployment claims. A review of studies conducted following the great recession of 2007 found that unemployment may contribute to hunger, the loss of housing, declines in birth weigh, increased symptom reporting, substance abuse, disability, medication use, and hospital visits, and untimely deaths due to suicide. (4) Meta-analyses have produced summary risks estimates of the effect of unemployment on several health outcomes. This analysis predicts the impact of Covid-19 unemployment on suicide and all cause mortality using these estimates.

## Methodology

Data on weekly unemployment claims came from the US Department of Labor. (5) Data on the age composition of the current labor force in 2019 came from the Bureau of Labor Statistics. (6) Data on all cause age specific death rates and age-adjusted suicide rates came from the National Bureau of Health Statistics. (7) The analysis used the average of the gender specific rates.

Estimates of suicide risk attributable to unemployment came from a meta-analysis of 16 studies. (8) Studies included in the meta-analysis were published after 1980, had a longitudinal cohort design, included data on duration of unemployment. Included studies came from Sweden, Finland, and Denmark, which are all countries with strong social safety nets and longitudinal employment and mortality registers. The overall pooled relative risk of suicide associated with long-term unemployment (average follow-up time 7.8 years) compared to those currently employed was 1.70 (95% CI 1.22 to 2.18). The authors found non-linear effects with the suicide hazard greater for unemployment < 5 years versus >5 years. Predictions applied in this analysis use estimates based on the 1-5 year follow up period (2.50; 95% CI 1.83 to 3.17). The publication did not provide gender and age specific risk estimates.

Estimates of age-specific excess all-cause mortality risk due to unemployment came from a published meta-analysis of 40 studies. (9) Studies were published between 1980 and 2008, came primarily from OECD countries, involved 20 million people, were of observational designs and had longitudinal follow up for up to 10 years. Overall pooled estimates of the hazard ratio in unemployed working aged people relative to the entire population were 1.63 (95% CI 1.49 to 1.79). This estimate represents the average effect over 10 years adjusted for age and other covariates, which varied by study. Estimates used for this analysis were age-group specific and adjusted for health risk covariates. The authors also estimated pooled hazard ratios stratified by duration of follow up but did not publish separate estimates for age and duration.

## Results

Applying age-specific hazard ratios from Milner et al. (2013) to the newly unemployed population distributed based on the composition of the current labor force and age-specific suicide rates, produces the results in the table below (Table 1). The central estimate based on the current number of the newly unemployed in the US population is approximately 9700 excess suicide deaths per year.

**Table 1.** Estimated excess annual suicides associated with U.S. Covid-19 pandemic unemployment.

| **Age Group** | **Employed Population (Millions)** | **Share of Employed Population** | **Number Unemployed After Covid-19 (Millions)** | **Age Specific US Suicide Rate** | **Estimated Excess Suicide Rate** | **Estimated Annual Excess Suicides** |
| --- | --- | --- | --- | --- | --- | --- |
| 16 to 24 | 19.32 | 12% | 4.42 | 14 | 21 | 944 |
| 25 to 34 | 35.81 | 23% | 8.18 | 18 | 27 | 2,197 |
| 35 to 44 | 33.13 | 21% | 7.57 | 18 | 27 | 2,033 |
| 45 to 54 | 32.04 | 20% | 7.32 | 20 | 31 | 2,241 |
| 55 to 64 | 26.89 | 17% | 6.15 | 20 | 31 | 1,881 |
| 65 to 74 | 8.44 | 5% | 1.93 | 17 | 26 | 492 |
| 75 + | 1.90 | 1% | 0.43 | 22 | 33 | 143 |
| **Totals** | **157.54** | **100%** | **36.00** |  |  | **9,786** |

Applying age-specific hazard ratios from Roelfs et al. (2011) to the newly unemployed population distributed based on the composition of the current labor force and age specific death rates, produces the results in the table below (Table 2). The central estimate based on the current number of the newly unemployed in the US population is approximately 66,000 excess deaths per year.

**Table 2.** Estimated excess annual deaths associated with U.S. Covid-19 pandemic unemployment.

| **Age Group** | **Employed Population (Millions)** | **Share of Employed Population** | **Unemployed After Covid-19 (Millions)** | **Age Specific US Death Rate (per 100,000)** | **Estimated Excess Death Rate (per 100,000)** | **Estimated Annual Excess Deaths** |
| --- | --- | --- | --- | --- | --- | --- |
| 16 to 24 | 19.32 | 12% | 4.42 | 72 | 53 | 2,321 |
| 25 to 34 | 35.81 | 23% | 8.18 | 130 | 95 | 7,765 |
| 35 to 44 | 33.13 | 21% | 7.57 | 195 | 150 | 11,366 |
| 45 to 54 | 32.04 | 20% | 7.32 | 398 | 306 | 22,439 |
| 55 to 64 | 26.89 | 17% | 6.15 | 886 | 222 | 13,612 |
| 65 to 74 | 8.44 | 5% | 1.93 | 1785 | 446 | 8,611 |
| 75 + | 1.90 | 1% | 0.43 | 4430 | 1107 | 4,816 |
| **Totals** | **157.54** | **100%** | **36.00** |  |  | **66,115** |

## Discussion

This analysis demonstrates the feasibility of quantifying indirect health impacts of societal responses to Covid-19. The risk assessment methodology, though simple, provides an evidence-based prediction of the impact of current unemployment since Covid-19 on excess suicide and pre-mature death.

This analysis is not a full accounting of all of the health impacts of either unemployment or the overall Covid-19 response. The choice of unemployment as a measure of health exposure and all-cause and suicide mortality as the measures of outcome is based on data availability. Unemployment is an earl indicator of the economic effects of Covid-19 responses. The consequences of unemployment on mortality and suicide are well studied and the subject of published meta-analyses. Further analyses may predict impact on unemployment on other outcomes, including health symptoms, disease conditions, and biological markers. (10–12)

The estimates have acknowledged uncertainties. For example, unemployment following Covid-19 may be short lived or prolonged. Some share of the predicted health effects might be mitigated by national policy countermeasures. Notably, most studies included the meta-analyses occurred in countries where social safety nets are robust.

Other economic effects, including the loss of income and health insurance, which are also associated with increased mortality, typically accompany unemployment. The effects of poverty on mortality are consistent across well designed longitudinal cohort studies. In the lower third of the income distribution each additional $10 000 of income reduces mortality risk by >50%. (13) Lack of health insurance is an independent predictor of mortality with smaller but consistent and statistically significant effects. (14) Given limited data on income and insurance effects and in the interest of not double counting impacts, these effects are not estimated separately.

Others have observed that overall mortality rates decline during economic downturns due to declines in traffic and possibly improvement in behavioral risk factors. Health impacts due to unemployment are distinct from and are experienced by a different group from the general population. How trade-offs are made between population benefits of economic decline and those directly impacted, such as the unemployed, is an important ethical question.

Accounting for the health benefits and costs of Covid-19 responses are important for optimizing public health. (15) In places where Covid-19 response has led to significant economic and social disruption, policy leaders might develop indicators and tools to monitor these effects. Longer-term income support, universal health insurance coverage and other policy interventions may be warranted to mitigate the indirect health impacts of Covid-19 response.

## Data Availability

All data used in this analysis is publicly available without a fee.

https://www.bls.gov/cps/cpsaat03.htm

https://www.cdc.gov/nchs/products/databriefs/db355.htm

## Contributions

Rajiv Bhatia designed, implemented, and wrote this analysis.

## Acknowledgments

Jennifer Hughes for her review of this manuscript.

## Conflicts of interest

None

## Funding

None

## Notes

### Competing Interest Statement

The authors have declared no competing interest.

### Clinical Trial

NA

### Funding Statement

The author received no funding or compensation for this work.

